# Atrial fibrillation begets atrial fibrillation – serum-driven amplification of atrial electrical instability

**DOI:** 10.64898/2026.10.09.26365177

**Authors:** Markus Bögner, Girish Ramesh, Pauline Fahjen, Judith Hüttemeister, Marzena Kirk, Emanuel Heil, Matthias Bock, Kun Zhang, Gabriele Boehm, Luis Eichelmann, Jin-Hong Gerds-Li, Ingo Hilgendorf, Volkmar Falk, Philipp Mertins, Gerhard Hindricks, Felix Hohendanner

**Affiliations:** Deutsches Herzzentrum der Charité (DHZC), Berlin, Germany; DZHK (German Centre for Cardiovascular Research), partner site Berlin; Max-Delbrück-Centrum für Molekulare Medizin (MDC)

**Keywords:** atrial fibrillation, serum, atrial cardiomyopathy, calcium signaling, circulating factors, proteomics

## Abstract

**Rationale:** Atrial fibrillation (AF) is sustained by electrical triggers and atrial substrate, but whether AF itself generates circulating factors that acutely reinforce arrhythmogenesis is unknown.

**Objective:** To determine whether rhythm-dependent serum bioactivity directly destabilizes atrial electrophysiology and to identify associated cellular mechanisms and circulating mediators.

**Methods and Results:** Cardiac samples were obtained from patients with persistent atrial fibrillation (AF) during AF and after restoration of sinus rhythm (SR) using electrical cardioversion. In isolated Langendorff-perfused mouse hearts, AF serum increased the duration, frequency, and cumulative burden of spontaneous atrial depolarizations compared with paired control perfusate. In human induced pluripotent stem cell derived atrial cardiomyocytes, AF patient serum decreased Ca²⁺-transient amplitudes without significant effect on kinetics or Ca²⁺ spark activity. Moreover, AF serum reduced peak sodium current without materially altering voltage-dependent gating or paced action-potential properties. Data-independent acquisition proteomics identified rhythm-associated changes in extracellular-matrix, proteolytic, inflammatory, and signaling proteins, including MMP9, Ccathepsin G, and 14-3-3 family members. Targeted pathway perturbation differentially modified serum-induced Ca²⁺ phenotypes, indicating context-dependent, multifactorial bioactivity. Serum-induced Ca²⁺ responses were not associated with indices of atrial disease severity, including low-voltage substrate burden and left atrial pressure.

**Conclusions:** Persistent AF is accompanied by rhythm-dependent circulating bioactivity that acutely promotes atrial electrical instability and Ca²⁺-dependent trigger formation. These findings extend the concept that “AF begets AF” beyond tissue-intrinsic remodeling and identify a potentially reversible serum-mediated feed-forward mechanism linking rhythm status and circulating proteomic signals in an atrial substrate severity independent fashion.

## Introduction

Atrial fibrillation (AF) is a self-reinforcing disease in which arrhythmogenic triggers interact with a progressively vulnerable atrial substrate. The seminal observation that sustained AF shortens atrial refractoriness and increases the stability of subsequent AF established the concept that “AF begets AF.”(1) Subsequent work extended this paradigm beyond electrical remodeling to fibrosis, inflammation, redox signaling, and disrupted intracellular Ca²⁺ homeostasis. In human atrial myocytes, enhanced diastolic sarcoplasmic-reticulum Ca²⁺ leak and increased electrogenic Na⁺/Ca²⁺ exchange promote delayed afterdepolarizations and triggered activity,(2) whereas oxidized Ca²⁺/calmodulin-dependent protein kinase II links redox stress to atrial proarrhythmia.(3) At the substrate level, fibrotic remodeling and low-voltage regions promote conduction heterogeneity and are associated with recurrent arrhythmia after rhythm-control interventions. (4, 5). The self-perpetuating nature of AF has nevertheless been considered predominantly a tissue-intrinsic process. AF is also accompanied by changes in circulating proteins related to myocardial stress, inflammation, extracellular-matrix turnover, proteolysis, and cellular signaling. Biomarker and proteomic studies have identified circulating signatures associated with prevalent and incident AF, (6, 7) and within-patient studies have shown that selected biomarkers change after restoration of sinus rhythm, indicating that at least part of the circulating phenotype is dynamically linked to rhythm status.(8) These observations have positioned circulating proteins as markers of AF and atrial disease severity; however, association does not establish biological activity. Whether the circulating milieu present during AF can itself acutely perturb atrial electrophysiology remains unknown. Establishing such a mechanism requires functional transfer experiments that separate the effects of circulating factors from the fixed structural and cellular abnormalities of the donor atrium. This distinction is difficult in conventional case control studies because circulating profiles are influenced by age, comorbidities, medication, hemodynamic load, and the extent of atrial cardiomyopathy. Paired sampling from the same patient during AF and after restoration of sinus rhythm reduces interindividual confounding and provides a strategy to identify rhythm-associated serum effects. Integrating this design with intact-organ electrophysiology, human atrial cardiomyocyte phenotyping, and unbiased molecular profiling can determine whether rhythm-dependent circulating signatures are merely correlates of AF or components of a feed-forward arrhythmogenic mechanism. We therefore tested the hypothesis that persistent AF generates a rhythm-dependent circulating bioactivity that directly promotes atrial electrical instability. Paired arterial and coronary-sinus serum samples were obtained from patients during persistent AF and after restoration of sinus rhythm. Serum bioactivity was assessed in Langendorff-perfused mouse hearts using multielectrode-array recordings and in human induced pluripotent stem cell–derived atrial cardiomyocytes by quantifying Ca²⁺ cycling, spontaneous diastolic Ca²⁺ release, sodium current, and action-potential properties. Data-independent acquisition proteomics was combined with targeted perturbation of candidate pathways, and serum-induced cellular phenotypes were related to electroanatomic and clinical measures of atrial cardiomyopathy. By linking paired human sampling to organ-level electrophysiology, cellular mechanisms, proteomic discovery, and substrate severity, this multiscale strategy was designed to determine whether the circulation is not merely a biomarker compartment but an active, potentially reversible component of the “AF begets AF” paradigm.

## Methods

### Study Design and Patient Population

Twenty-three patients with persistent AF undergoing pulmonary vein isolation at the Deutsches Herzzentrum der Charité were included. All patients were in AF at the beginning of the procedure and underwent electrical cardioversion during the intervention. The study was approved by the institutional ethics committee of Charité–Universitätsmedizin Berlin under approval number EA4/166/21 and was conducted in accordance with the Declaration of Helsinki. All participants provided written informed consent.

Procedural anesthesia was initiated at the beginning of the pulmonary vein isolation procedure according to the institutional protocol and maintained during both sampling time points. Clinical, echocardiographic, laboratory, and procedural data were obtained from the medical record. Serum samples were coded before experimental use. Experiments and analyses were performed by investigators blinded to patient identity and rhythm condition. Experimental allocation and the order of experimental conditions were not randomized.

### Blood Sampling and Serum Preparation

Blood was collected from the left atrium (“arterial”) and the great cardiac vein (“coronary sinus”) using angiographic and ultrasound-guidance respectively, while patients were in persistent AF (Figure 1A). Electrical cardioversion was subsequently performed, and paired arterial and coronary-sinus samples were obtained >15 minutes after restoration of SR. Rhythm status was confirmed by surface electrocardiography and intracardiac electrograms.

**Figure 1.**
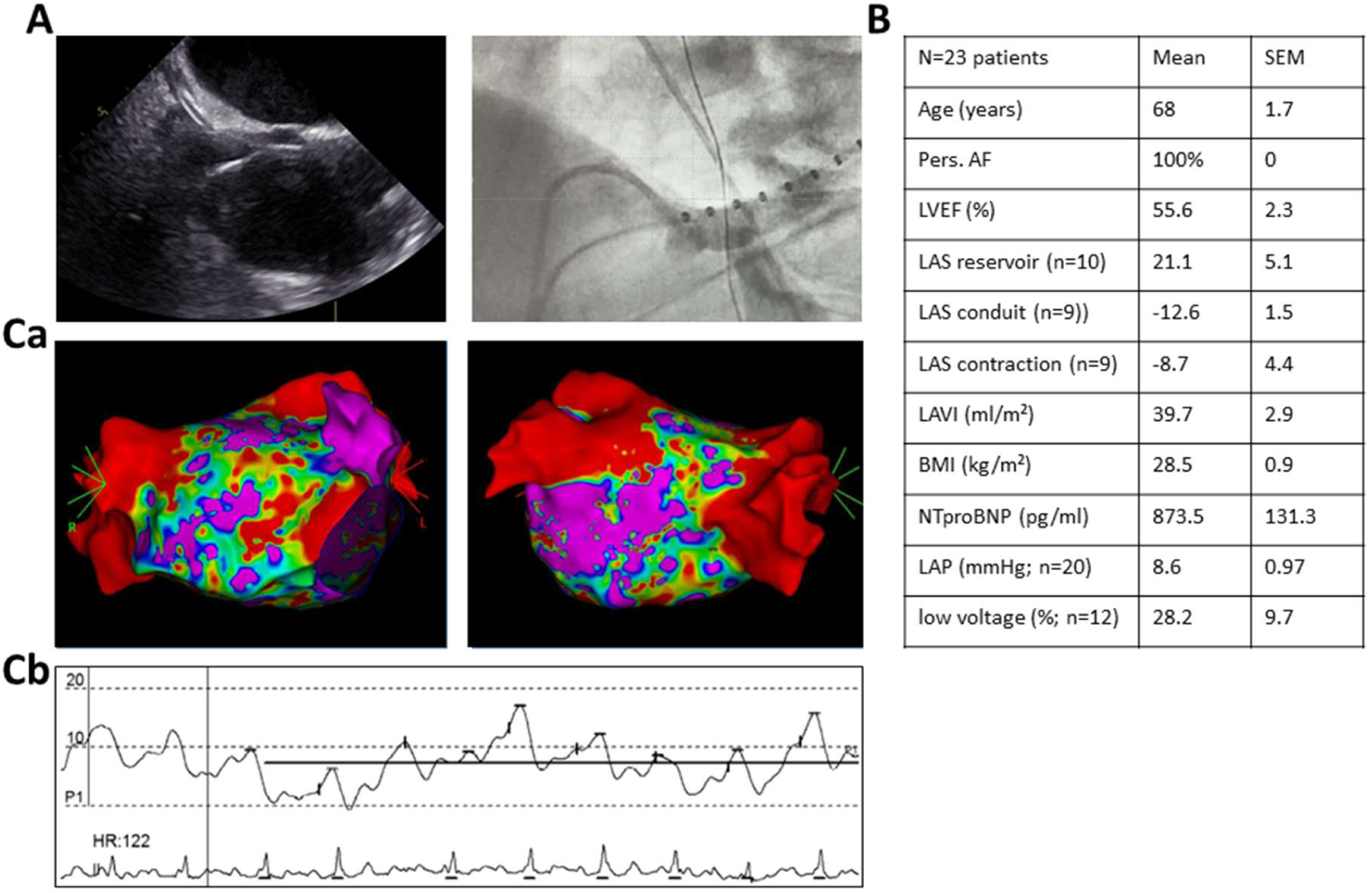
Multimodal characterization of the left atrial substrate in persistent atrial fibrillation. (A) Representative echocardiographic image (left), Fluoroscopic visualization of intracardiac catheter positioning during the electrophysiological procedure (right). (B) Clinical, echocardiographic, laboratory, and invasive characteristics of the 23 patients with persistent atrial fibrillation. Data are presented as mean±SEM unless otherwise indicated. Sample sizes are specified for variables not available in the entire cohort. (Ca) Three-dimensional left atrial bipolar voltage maps in complementary views obtained using the CARTO 3 system and a multipolar mapping catheter. Low-voltage myocardium was defined as a bipolar electrogram amplitude <0.5 mV. (Cb) Representative invasive left atrial pressure recording with simultaneous electrocardiographic tracing.

Blood was centrifuged according to a standardized protocol. Serum was separated, divided into aliquots, and stored at −80°C until use. Paired AF and SR samples (final concentration of 16.7%) (9) from the same patient were processed identically and, whenever possible, studied within the same experimental series. Partially overlapping subsets of patient-derived serum samples were used for functional characterization, comprising six patients for automated patch-clamp analysis, seven for Ca²⁺-transient imaging, and nine for Langendorff/MEA experiments. Availability of serum varied from patient to patient, leading to different amounts available for the study assays: Overall, serum samples from 11 unique patients were analyzed across the functional assays, including three patients whose samples were tested in all functional assays. Paired AF–SR proteomic analyses included 62 serum samples from 16 patients. Fifteen patients contributed coronary sinus and arterial samples collected during both AF and SR, whereas one patient contributed only coronary sinus samples collected during both rhythm states.

### Electroanatomic Mapping

Three-dimensional left atrial electroanatomic mapping was performed using the CARTO 3 system and a multipolar mapping catheter according to the institutional high-density mapping protocol (Pentaray catheter, Biosense Webster). Mapping points were accepted according to local standards for signal quality and catheter position. Low-voltage myocardium was defined as a bipolar electrogram amplitude of <0.5 mV. Low-voltage area was quantified as the absolute area and as the proportion of the mapped left atrial surface (map obtained during sinus rhythm). Low-voltage measurements were available in 12 patients. Left atrial pressure was measured invasively in 20 patients according to the institutional procedural protocol (Figure 1C)

### Isolated-Heart Electrophysiology and Multielectrode-Array Recording

Animal experiments were approved by the responsible regulatory authority and were conducted in accordance with institutional and national guidelines (2023-019-T). Male C57BL/6N wild-type mice aged 25-32 weeks were used for the study.

Mouse hearts were rapidly excised, cannulated through the aorta, and mounted on a modified Langendorff apparatus. Hearts were retrogradely perfused with physiological solution under temperature-controlled conditions. After stabilization, atrial field potentials were recorded using a multielectrode-array (MEA) system (Multichannel Systems).

Each heart was sequentially exposed to control perfusate, SR serum, and paired AF serum (1:6) from the same patient for a period of 1 minute prior to the measurements at a flowrate of 1.1mL/min and washout was carried out using control perfusate (in mmol/L: 135 NaCl, 4.7 KCl, 0.6 KH₂PO₄, 0.6 Na₂HPO₄, 1.2 MgSO₄·7H₂O, 10 HEPES, 30 Taurine, 10 Glucose, and 0.01 Blebbistatin) for a period of 1-2 minutes at the same flow rate as serum. Serum exposures were separated by washout with control perfusate. The exposure sequence was fixed and was not randomized. All the experiments were carried out at 37 °C. Fourteen hearts were studied using CS serum from 9 independent patients.

Atrial electrophysiological responses were assessed using an external burst-pacing protocol during MEA recordings by placing an EcoFLEX36 MEA, containing 64 electrodes on the left atrium. Biphasic rectangular stimuli (±10 mV, 2 ms total duration; 1 ms positive and 1 ms negative phase) were delivered using an external stimulator. The protocol consisted of 1 min pacing at 100 bpm (1.67 Hz), followed by a 4 min unstimulated baseline recording. Subsequently, four 30 s burst-pacing periods alternating between 500 bpm (8.33 Hz) and 1000 bpm (16.67 Hz) were applied, with each burst followed by a 4 min unstimulated recording period. MEA signals were acquired using the Cardio2D software (Multichannel Systems). The total duration of the entire protocol was 23 min. Spontaneous depolarizations were detected using a baseline-dependent amplitude threshold because field-potential amplitudes differed among recordings. A spontaneous depolarization was defined as at least 2 suprathreshold excitation peaks occurring within a 0.5 second interval between 2 consecutive regularly conducted atrial complexes. An event was considered terminated when a stable baseline without additional suprathreshold activity persisted for ≥0.5 seconds.

The quantified outcomes were event duration, event frequency expressed as events per minute, the occurrence of prolonged events lasting >3 seconds, and cumulative spontaneous depolarization burden.

Depolarization burden was calculated by combining AF event frequency with a duration-weighted measure of AF events. AF events were categorized according to their duration (1–3 s, 3–10 s, and >10 s), with weighting factors of 1, 3, and 10, respectively. AF episode frequency was calculated as the total number of AF events divided by the total recording duration in minutes. The depolarization burden was then calculated as the product of the duration-weighted AF event measure and AF episode frequency.

### Generation of Human iPSC-Derived Atrial Cardiomyocytes

Human iPSC-aCMs were differentiated from the BIHi0005A control iPSC line derived from a healthy male donor. Atrial cardiomyocyte differentiation was performed using temporal modulation of canonical Wnt signaling followed by retinoic-acid–directed atrial specification, based on the protocol described by Cyganek et al.(10)

Briefly, iPSCs were expanded in Essential 8 medium until approximately 80% confluent. Cardiac mesoderm formation was initiated on day 0 by transient activation of Wnt signaling using CHIR-99021 (Sigma, Cat#SML1046) in RPMI medium without insulin. Wnt signaling was subsequently inhibited using IWP2 (Sigma, Cat#681671) on day 3. Atrial lineage specification was induced at day 4 by exposure to 1 µmol/L retinoic acid for 72 hours.

Following cardiac specification, cells were maintained in RPMI 1640 supplemented with B27 containing insulin. Spontaneous contractions were generally observed from day 9-12. Lactate-based metabolic selection was performed on day 15 to enrich the cardiomyocyte population. Cardiomyocytes aged 90±20 days were used for functional studies. The serum-exposure and confocal Ca²⁺-imaging procedures were adapted from the previously published workflow of Dimai et al.(9)

### Confocal Ca²⁺ Imaging

iPSC-aCMs were loaded with 1.8 µmol/L Fluo-4 AM for 30 minutes at 37°C and 5% CO₂. Cells were subsequently washed with Tyrode solution containing, in mmol/L: 135 NaCl, 4.7 KCl, 0.6 KH₂PO₄, 0.6 Na₂HPO₄, 1.2 MgSO₄·7H₂O, 10 HEPES, 30 taurine, 10 glucose, and 1.8 CaCl₂.

Cardiomyocytes loaded with Fluo-4 AM were exposed to control conditions, SR serum, or AF serum (1:3, 33% concentration) for 30 minutes, and then imaged in 16.7% serum concentration. SR and AF sera were paired at the patient level, but measurements were obtained from distinct cells rather than sequentially from the same cardiomyocyte.

Confocal line-scan images were acquired at 512 lines per second using a ×40 oil-immersion objective. The scan line was positioned along the cardiomyocyte axis with a spatial resolution of 0.12 µm per pixel. Cells were electrically field stimulated at 1 Hz for 10 seconds, followed by a 10-second period without stimulation to assess spontaneous diastolic Ca²⁺ release.

Fluorescence was expressed as F/F₀, where F represented background-corrected fluorescence and F₀ represented background-corrected diastolic fluorescence during paced stimulation. The analyzed parameters were Ca²⁺-transient amplitude, time to peak, time to 50% release, monoexponential decay time constant τ, and spontaneous Ca²⁺-spark frequency.

Ca²⁺ sparks were detected using the Spark Master plugin for ImageJ. Spark frequency was normalized to scan length and recording duration and expressed as sparks per 100 µm per second. Seven independent paired patient-serum samples were included in the Ca²⁺-imaging experiments.

### Candidate-Pathway Perturbation

Proteomics-guided perturbation experiments were performed to examine the contribution of MMP9, 14-3-3 signaling, and Cathepsin G to serum-induced Ca²⁺ abnormalities. iPSC-aCMs were exposed to SR or AF serum alone or in the presence of MMP9 inhibitor II (10µM, Sigma #444249), the 14-3-3 interaction inhibitor Difopein (5µM, Adooq Bioscience #A14917), or a Cathepsin G inhibitor (5µM, Thermoscientific #J65957.MCR) diluted in Tyrode’s solution for 30 minutes before imaging. Separate cells were used for the individual inhibitor conditions. Ca²⁺-transient kinetics and spontaneous Ca²⁺-spark frequency were quantified as described above and compared with the corresponding serum-only condition.

### Automated Patch-Clamp Electrophysiology

For patch-clamp experiments, iPSC-aCMs were exposed to control conditions, SR serum, or paired AF serum for 1 hour before preparation of a single-cell suspension. Separate cells were studied under each experimental condition.

Automated whole-cell patch-clamp recordings were performed using a QPatch 48 system (Sophion Bioscience). The extracellular solution contained, in mmol/L: 145 NaCl, 4 KCl, 2 CaCl₂, 1 MgCl₂, 10 HEPES, and 10 glucose and was adjusted to pH 7.4 with NaOH and to 290–310 mOsm. The intracellular solution contained, in mmol/L: 120 KCl, 31.25 KOH, 10 EGTA, 10 HEPES, 5.374 CaCl₂, 1.75 MgCl₂, and 4 Na₂-ATP and was adjusted to pH 7.2 with KOH and to 285–290 mOsm.

Peak sodium current was elicited using a 300-ms depolarizing step from a holding potential of −90 mV to −30 mV. Sodium current–voltage relationships were obtained using 10-ms voltage pulse from −90 to +70 mV in 10-mV increments from a holding potential of −90 mV. Currents were normalized to membrane capacitance and expressed as current density. Reversal potential, half-maximal activation voltage, and maximum conductance were derived from Boltzmann fits. Integrated sodium current was calculated over the voltage range from −40 to +10 mV.

Action potentials were recorded in current-clamp mode using 2-ms, 1-nA current injections delivered at 1-Hz intervals. Threshold potential, peak potential, maximum diastolic potential, action-potential amplitude, and action-potential duration at 50% and 90% repolarization were quantified.

Data were sampled at 25 kHz and low pass filter using a 5 kHz Bessel filter. Fast R_s_ compensation was applied and maintained at 80% during sodium current measurements. Recordings were included when membrane resistance was >100 MΩ and membrane capacitance was >5 pF. Leak subtraction was executed online using QPatch software using linear leak to isolate ionic currents from the background leak. Six independent paired patient-serum samples were included.

### Serum Proteomics

Proteomic profiling was performed in arterial and coronary-sinus serum samples. Sample preparation was automated using an Agilent BRAVO system. Serum was diluted 1:10 with water, mixed with lysis buffer (2% SDC, 20 mM DTT, 80 mM CAA, 22 mM Tris, pH 8), and heated at 95°C for 10 min. Proteins were digested overnight at 37°C with 4 µg each of trypsin (Promega) and LysC (Wako) in 50 mM HEPES (pH 8). Acidification and SDC precipitation were achieved with 10% FA, followed by dilution with 1% FA. After centrifugation at 1200g for 10 min, the supernatant was desalted on C18 cartridges, eluted in 50% ACN/0.1% FA, dried, and stored at −20°C.

For analysis, peptides were reconstituted in 3% ACN/0.1% FA, and 1 µg was injected into a Vanquish Neo HPLC system coupled to an Orbitrap Exploris 480 mass spectrometer. Peptides were separated using a 32-minute gradient with a flow rate of 250 nL/min on a 20 cm reverse-phase column packed in house with 1.9 µm C18 beads (ReproSil Pur, Dr. Maisch GmbH) with an inner diameter of 75µm. The concentration of Buffer B (90% ACN/0.1% FA) was increased from 2% to 7% in the first minute, followed by a gradual rise to 20% over 19 minutes, then to 30% over the next 9 minutes, and finally to 60% in the last 3 minutes. MS data were acquired in data-independent acquisition (DIA) mode. Full scans were performed at a resolution of 120K with a normalized AGC target of 300%, a mass range of 350-1650 m/z, and a maximum injection time of 20 ms. Each full scan was followed by 40 MS2 scans with variable window widths, a resolution of 30K, a normalized AGC target of 3000%, stepped collision energy settings (26, 29, 32), and a maximum injection time of 35 ms. The cycle time was set to 3 seconds.

Protein and peptide identification was performed in Spectronaut (version 18.2) using directDIA and the Uniprot database including isoforms (2022–03). Default settings were kept, and Carbamidomethyl (C) was set as fixed modification and Acetyl (Protein N-term), Deamidation (NQ), and Oxidation (M) as variable modifications.

A total of 719 protein groups were identified. Potential protein contaminants were removed and log_2_-transformed protein expression values were filtered to exclude proteins with < 80% valid values, resulting in 504 proteins entering statistical analysis. Protein intensities were median normalized and missing values were imputed from a normal distribution (width = 0.3 SD, downshift = 1.8). Differential abundance was assessed using a linear model implemented in limma (11) on log2-transformed protein intensities. SR versus AF was assessed separately for arterial (A) and coronary sinus (CS) samples, as well as in a combined contrast across both sampling locations. For the combined comparison, the A- and CS-specific coefficients were given equal weight, such that each sampling location contributed 50% to the contrast. Patient identity was included as a blocking factor to account for the correlation between repeated measurements from the same patient, using ‘duplicate Correlation ()’. The AF–SR analysis included 62 serum samples from 16 patients: 15 patients had samples from both sampling locations in both AF and SR (four samples per patient), whereas one patient had only CS samples in both conditions.

Empirical Bayes variance moderation with intensity-dependent variance modeling (‘trend = TRUÈ) was applied. P-values were adjusted for multiple testing using the Benjamini–Hochberg method, with an FDR threshold of 0.1.

### Peptide specificity and preanalytical sensitivity analyses

For isoform determination of YWHAE, identified peptide sequences were inspected to determine whether they were specific to the canonical YWHAE isoform (UniProt P62258) or the short transcript ENST00000496706. The dataset was additionally screened for detection of the seven human 14-3-3 paralogues YWHAE, YWHAZ, YWHAH, YWHAB, YWHAQ, YWHAG, and SFN.

Potential sample preprocessing effects were assessed at the protein level using median-normalized, non-imputed log_2_ protein intensities. For each patient and sampling location, paired changes were calculated as log_2_ protein abundance during AF minus log_2_ abundance after restoration of SR. Changes in YWHAE, YWHAZ, MMP9, and CTSG were correlated with corresponding changes in the platelet-associated proteins PF4V1 and PPBP and the neutrophil-associated proteins MPO and LTF. Correlations among YWHAE, YWHAZ, MMP9, and CTSG were assessed in the same way. The analyses included only pairs with complete measurements at both time points. Spearman rank correlation coefficients and nominal P values were reported. As these analyses were exploratory, no correction for multiple testing was applied.

For visualization of broader platelet- and neutrophil-associated protein patterns, a heatmap was generated using YWHAE, YWHAZ, the manually selected platelet-associated proteins PPBP, TLN1, THBS1, PFN1, MMRN1, FLNA, and VCL, and the neutrophil-associated proteins MPO, LCN2, and LTF. Normalized log_2_ protein abundances were standardized separately for each protein across samples, and z-scores were truncated at −2.5 and +2.5 for visualization. Arterial and coronary-sinus samples were displayed separately, with AF and SR samples ordered by patient to preserve the paired structure. Samples were not clustered, whereas proteins were hierarchically clustered within the predefined platelet, neutrophil, and 14-3-3 protein groups.

### Statistical Analysis

Statistical analyses were performed using SPSS and GraphPad Prism version 10. All tests were 2-sided, and P<0.05 was considered statistically significant. Data distribution was assessed using the Shapiro–Wilk test.

For isolated-heart experiments, the heart was considered the experimental unit. Measurements obtained under control, SR-serum, and AF-serum conditions in the same heart were analyzed using repeated-measures one-way ANOVA followed by multiplicity-adjusted post hoc comparisons. When only two conditions were compared, a paired Student t test was used.

For cellular experiments, individual cardiomyocytes constituted subsamples, whereas the independent patient-serum sample was considered the biological replicate. Measurements from cells exposed to the same patient serum under the same condition were therefore averaged to generate one patient-specific value per condition. Paired SR- and AF-serum conditions were compared using paired Student t tests or Wilcoxon matched-pairs signed-rank tests when normality assumptions were not fulfilled.

Candidate-inhibitor experiments were analyzed using patient-specific mean values. Serum-only and inhibitor conditions generated using serum from the same patient were compared using repeated-measures one-way ANOVA followed by multiplicity-adjusted post hoc comparisons. The Friedman test with Dunn multiple-comparison testing was used when parametric assumptions were not met.

Associations between serum-induced cellular responses and clinical or electroanatomic variables were assessed using patient-specific mean cellular values and Spearman rank correlation. Simple linear-regression lines were included for visualization.

Differential protein abundance was analyzed using the paired multilevel limma workflow described above.

Data are presented as mean±SD or mean±SEM, as specified in the corresponding figure legends. Individual cell measurements may be displayed to illustrate the distribution of cellular responses; inferential statistical testing was performed using patient-specific mean values. The numbers of patients, serum samples, hearts, and cells included in each analysis are reported in the figure legends.

## Results

### Patient characteristics and electroanatomic substrate

A total of 23 patients undergoing rhythm control procedures were included in the clinical cohort. All patients exhibited persistent AF at the time of serum sampling. Mean age was 68 ± 1.7 years, with preserved left ventricular systolic function (LVEF 56 ± 2%). Left atrial remodeling was evident, as reflected by an increased left atrial volume index (LAVI 39.7 ± 2.9 ml/m²) and a substantial burden of low-voltage areas on three-dimensional electroanatomic mapping (28.2 ± 9.7% in patients with available voltage data). NT-proBNP levels were markedly elevated (873.5 ± 131.3 pg/ml), consistent with advanced atrial disease. Detailed clinical and mapping characteristics are summarized in Figure 1.

### AF serum induces spontaneous atrial depolarizations in Langendorff-perfused mouse hearts

To determine whether patient serum exerted direct electrophysiological effects at the intact-organ level, 14 isolated mouse hearts were studied using paired SR and AF serum samples obtained from nine patients. Each heart was sequentially exposed to control perfusate (Tyrode solution), SR serum, and AF serum from the same patient pair. Thus, each heart was tested with one patient pair, and some patient pairs were tested in more than one mouse heart where applicable. Spontaneous atrial depolarizations were uncommon during control perfusion, with a mean event duration of 15±29 seconds, an event frequency of 0.28±0.50 events/min, and a cumulative depolarization-burden index of 17±35. Exposure to SR serum was associated with an event duration of 21±33 seconds, an event frequency of 0.44±0.63 events/min, and a depolarization burden of 29±59. AF-serum exposure produced the highest values across these outcomes, with an event duration of 96±162 seconds, an event frequency of 1.10±1.48 events/min, and a cumulative burden of 341±718. Compared with control perfusate, AF serum significantly increased spontaneous depolarization duration, event frequency, and cumulative depolarization burden (P<0.05; Figure 2). The frequency of prolonged events lasting >3 seconds was numerically higher during AF-serum exposure. SR-serum values were intermediate between control and AF-serum conditions; however, no significant differences were detected between SR serum and either control or AF Serum.

**Figure 2.**
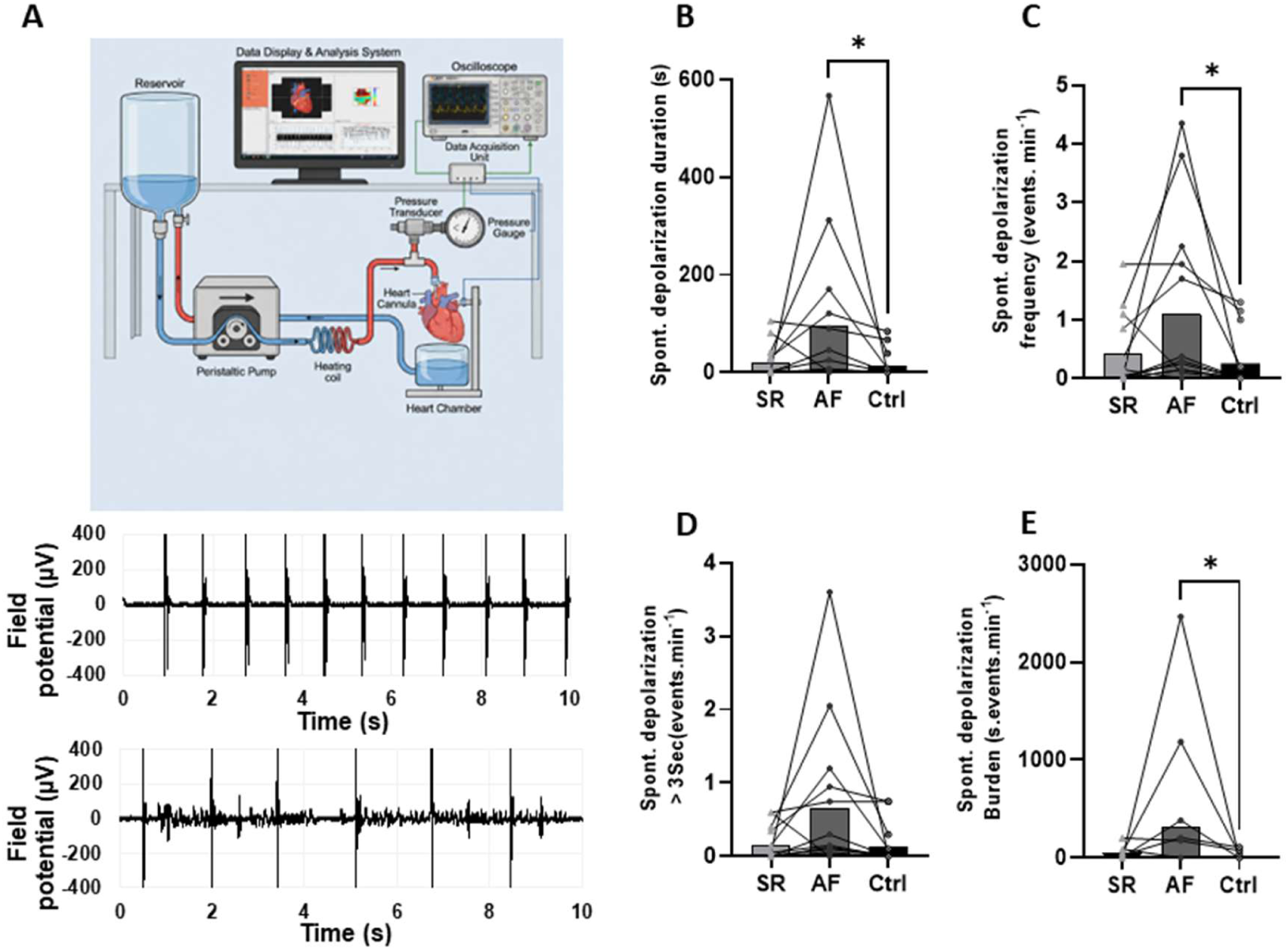
Patient serum induces spontaneous depolarization in Langendorff-perfused mouse atria assessed by MEA. **(A)** Schematic of the experimental setup. **(B–E)** Quantification of spontaneous depolarization activity (*n* = 14 hearts; 9 independent patient serum samples). **(B)** Duration of spontaneous depolarizations (s). **(C)** Spontaneous depolarization frequency (events·min⁻¹). **(D)** Prolonged spontaneous depolarization events (>3 s·min⁻¹). **(E)** Spontaneous depolarization burden (s. events.min^-1^). \**p*<0.05. Individual data points represent paired measurements from the same heart across conditions; connecting lines indicate pairing. Bars show mean. Statistical comparisons were performed using Kruskal-Wallis test; p-values <0.05 are considered statistically significant.

### AF Serum Produces Selective Changes in Sodium-Current Properties Without Reducing Integrated or Peak Current

We next examined fast sodium current (I_Na_) properties in iPSC-aCMs following exposure to SR or AF patient sera for six patient serum samples using an automated patch clamp technique. Representative whole-cell recordings demonstrated a robust, rapidly activating and inactivating inward current consistent with I_Na_ (Figure 3A). Quantification of peak I_Na_ amplitude revealed a significant reduction in cells treated with patient sera compared with control, with the most pronounced decrease observed in AF (Control: −2883±1662; SR: −2417±1382; AF: - 2374±1427pA) (Figure 3 H). This was further supported by current–voltage (I–V) relationships, which showed reduced current density across a range of test potentials in SR and AF conditions, reaching statistical significance at voltages near peak activation (approximately −40 to −10 mV) (Figure 3 B). To determine whether these changes were accompanied by alterations in channel gating, we analyzed voltage dependence of activation through transformed Boltzmann fit from the Na_v_ IV measurements. The reversal potential (V_rev_) was significantly increased in AF compared with SR (Control: 71±12; SR: 72±8; AF: 77±11mV) (Figure 3D), whereas the half-maximal voltage of activation/inactivation (V_1/2_) was not significantly different between groups (Control: −48±7; SR: −47±6; AF: −48± 8mV) (Figure 3 E). Similarly, maximal conductance (G_max_) showed no statistically significant differences across Ctrl, SR, and AF conditions (Control: 28±16; SR: 23±13; AF: 25±17mV) (Figure 3 F), despite a trend toward reduced values in patient serum-treated iPSC-aCMs. Collectively, these data indicate that exposure to AF patient serum reduces peak sodium current density in iPSC-aCMs without major shifts in voltage-dependent gating, suggesting that decreased channel availability, rather than altered biophysical properties, underlies the observed electrophysiological remodeling.

**Figure 3.**
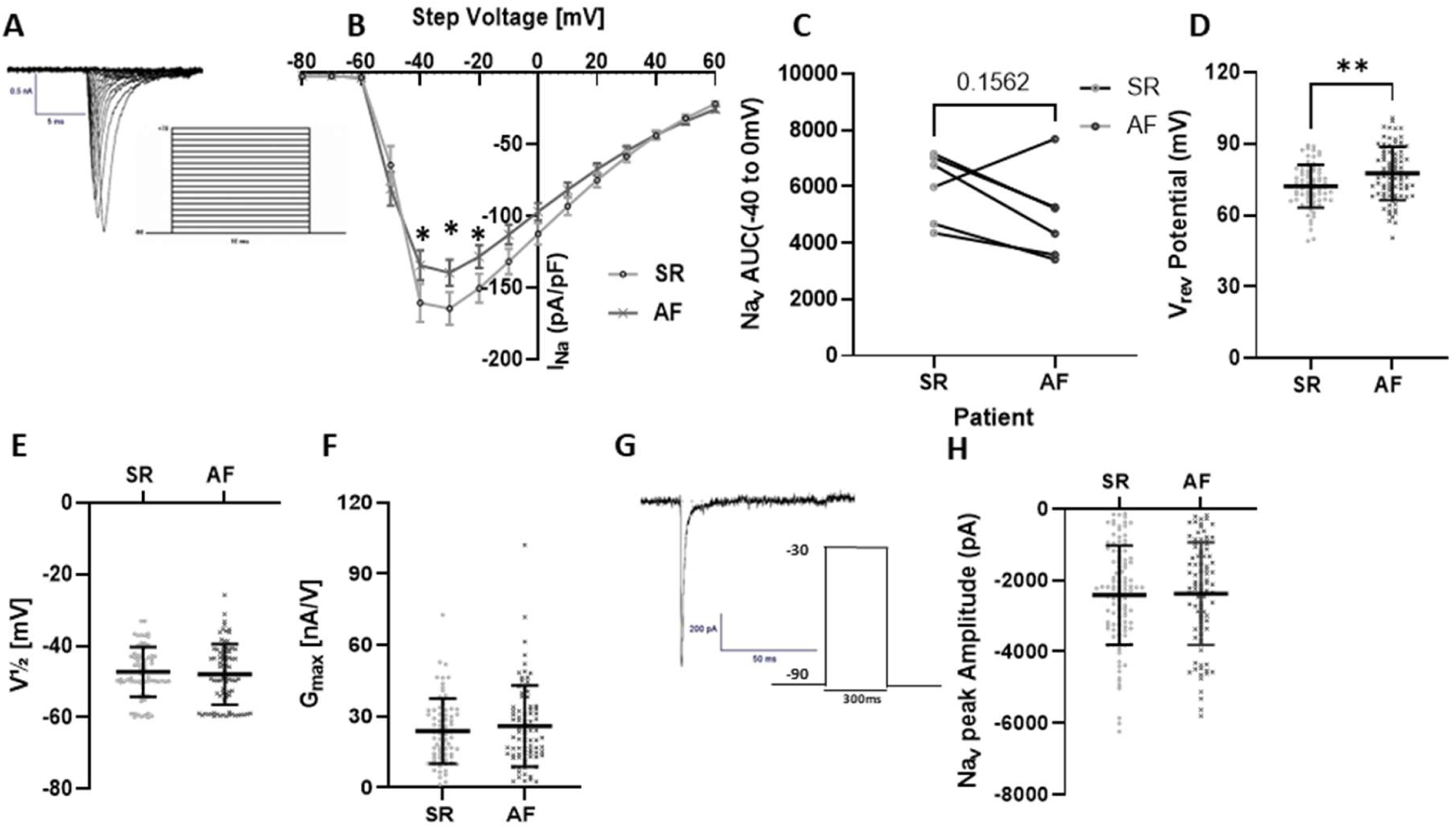
Modulation of peak sodium current (I_Na_) by patient serum in cardiomyocytes. **(A)** Representative I–V relationship of I_Na_ elicited by depolarizing steps from −90 to +70 mV (10 mV increments, 10 ms duration). **(B)**Mean current–voltage relationship expressed as current density (pA·pF⁻¹) (6 patient sera; Control: *n* = 89 cells; SR: *n* = 73; AF: *n*= 91). **(C)** Area under the Nav IV (−40 to 0 mV) calculated from patient-averaged current density values following exposure to paired SR and AF serum samples (n = 6). Wilcoxon matched-pairs signed-rank test, P = 0.156). **(D–F)** Steady-state activation parameters derived from Boltzmann fits. **(D** Reversal potential (V_rev_). **(E)** Half-maximal activation voltage (V_1/2_). **(F)** Maximum conductance (G_max_). **(G)** Representative inward sodium current (I_Na_) recorded at −30 mV. Inset shows the voltage protocol. **(H)** Quantification of peak I_Na_ amplitude. Data derived from 6 independent patient serum samples (Control: *n* = 95 cells; SR: *n* = 104; AF: *n* = 93). Individual data points represent single cells; bars indicate mean ± SEM. Statistical comparisons were performed using paired Student’s t-test; p-value<0.05 considered statistically significant.

### Patient Serum Does Not Materially Alter Paced Action-Potential Properties

Action potentials were recorded during 1-Hz pacing in iPSC-aCMs exposed to control conditions or serum from six patients. No statistically significant differences were identified in threshold potential, peak potential, maximum diastolic potential, action-potential duration at 90% repolarization, or action-potential amplitude among control, SR-serum, and AF-serum conditions (Figure 4). Action-potential duration at 50% repolarization was numerically shorter following AF-serum exposure than following SR-serum exposure, but this difference did not reach statistical significance (P=0.0592). These findings indicate that the limited changes in sodium-current properties were not accompanied by major alterations in paced action-potential morphology under the recording conditions used.

**Figure 4.**
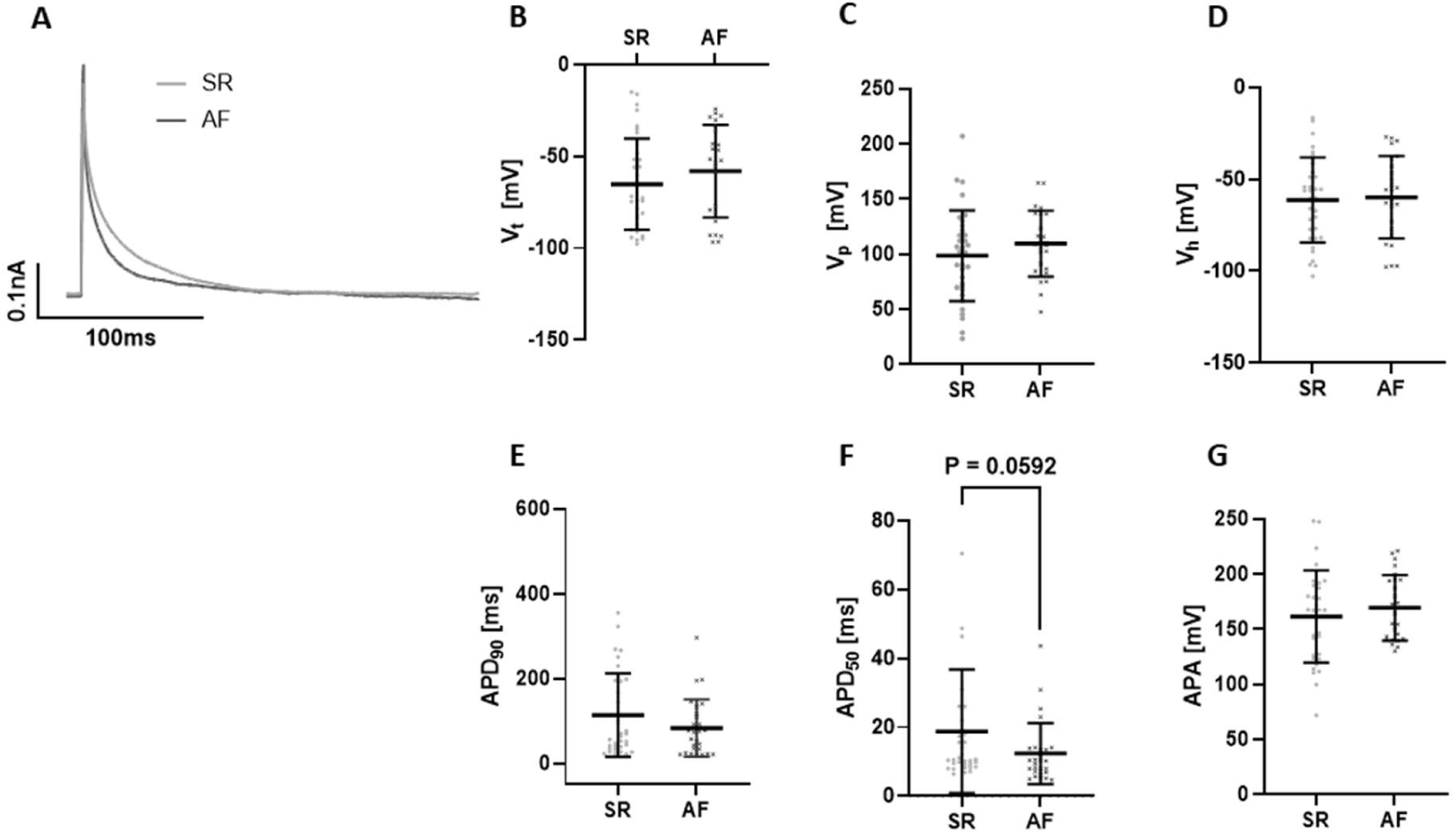
Effects of patient serum on action potential properties in iPSC-derived atrial cardiomyocytes. **(A)** Representative action potential (AP) traces recorded from iPSC-derived atrial cardiomyocytes: treated with SR Serum (light grey) and AF serum (dark grey). **(B–G)** Quantitative analysis of AP parameters obtained from paced recordings (6 independent patient serum samples; Control: *n* = 28 cells; SR: *n* = 34; AF: *n* = 25). **(B)** Average Threshold potential (Vt). **(C)** Average Peak potential (Vp). **(D)** Average Maximum diastolic (hyperpolarization) potential (Vh). **(E)** Mean Action potential duration at 90% repolarization (APD90). **(F)** Mean Action potential duration at 50% repolarization (APD50). **(F)** Action potential amplitude (APA). Individual data points represent single cells; bars indicate mean ± SEM. Statistical comparisons were performed using paired Student’s *t*-test; p<0.05 are considered statistically significant.

### Proteomic Profiling Identifies Rhythm-Associated Candidate Proteins

Proteomic profiling was performed in 62 serum samples obtained during AF and after restoration of SR from 16 patients. Differential-abundance analysis revealed partially distinct protein signatures depending on the sampling compartment (Figure 5A–B). In arterial serum, several proteins showed greater abundance in the AF-associated direction, with HBE1 and YWHAZ displaying the strongest differences, while CA2 and LCAT also exceeded the predefined significance threshold (Figure 5A). YWHAE, MMP9, and cathepsin G (CTSG) showed more modest changes. In coronary-sinus serum, fewer proteins demonstrated pronounced rhythm-associated differences, with TUBB representing the strongest signal, whereas MMP9, YWHAE, YWHAZ, and CTSG showed smaller changes that did not reach the same statistical threshold (Figure 5B). Principal-component analysis demonstrated interindividual and sample-compartment heterogeneity, with incomplete separation between AF and SR samples (Figure 5C & D). Nevertheless, paired differential-abundance analyses identified a set of rhythm-associated candidate proteins (adjusted p-value < 0.1). Proteins prioritized for functional follow-up included matrix metalloproteinase-9 (MMP9) and the 14-3-3 family members YWHAE and YWHAZ. Cathepsin G was included as an additional proteolytic candidate for pathway-interference experiments. The proteomic data therefore nominated extracellular-matrix, proteolytic, and intracellular signaling pathways potentially associated with serum collected during AF, but did not identify a single dominant candidate capable of explaining the complete functional phenotype.

**Figure 5.**
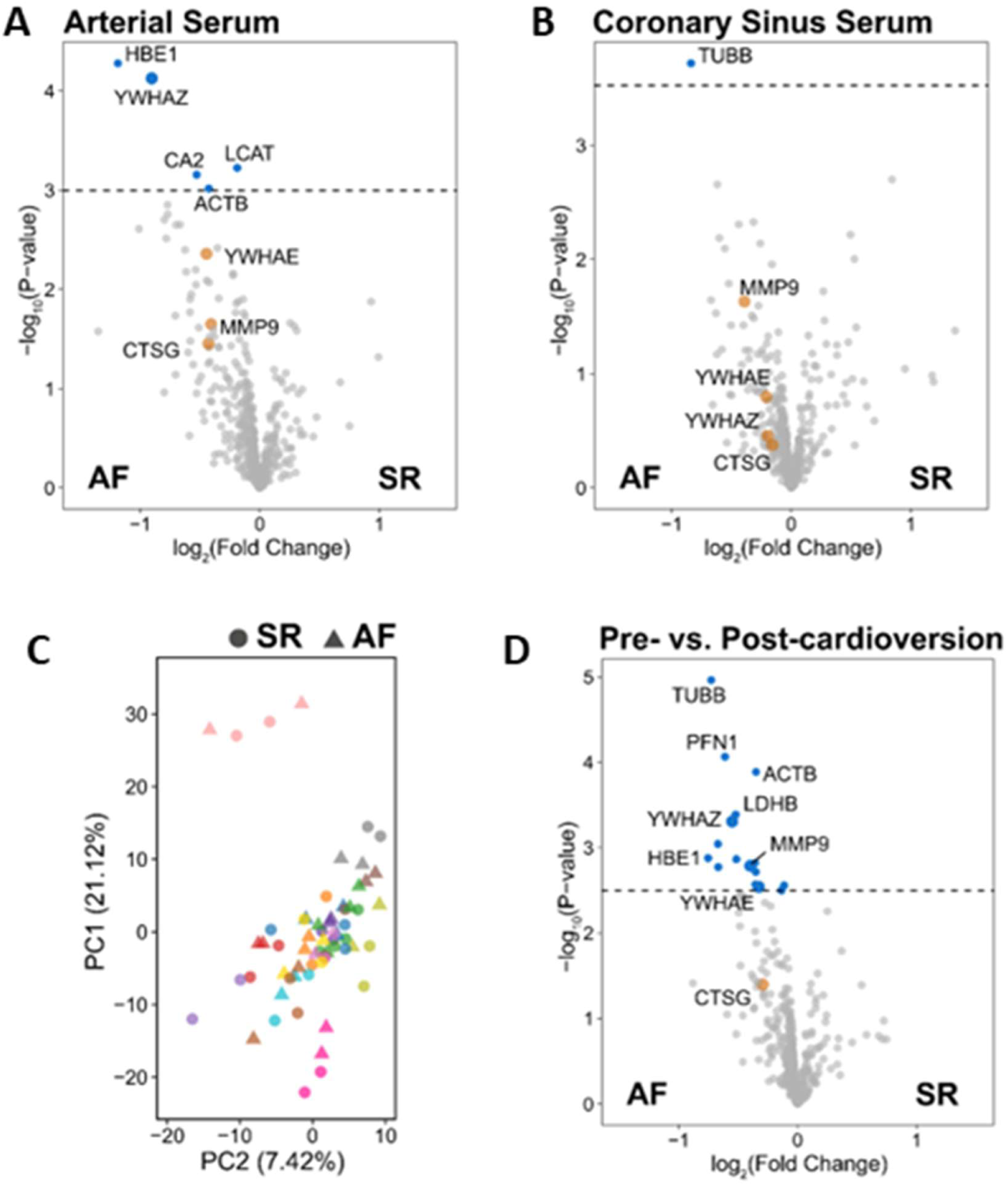
Differential proteomic signatures in arterial and coronary sinus serum from patients with atrial fibrillation. **(A)** Volcano plot showing differential protein abundance between atrial fibrillation (AF) and sinus rhythm (SR) in the arterial serum (n = 15 patients). **(B)** Volcano plot showing differential protein abundance between atrial fibrillation (AF) and sinus rhythm (SR) in the coronary sinus serum (n = 16 patients). **(C)** Principal component analysis (PCA) of proteomic profiles from arterial and coronary sinus samples. Each point represents an individual patient sample, coloured by patient; AF and SR groups are indicated by point shape. Variance explained by each principal component (PC1 / PC2) is shown on the axes. **(D)** Volcano plot showing differential protein abundance between AF and SR, using a combined contrast across arterial and coronary sinus samples, with equal weight given to each sampling location. Proteins with a Benjamini–Hochberg adjusted p-value < 0.1 are highlighted in blue (n = 16 patients).

### Peptide specificity and preanalytical sensitivity analyses

Among the seven 14-3-3 paralogues, only YWHAE and YWHAZ were quantified in more than 80% of samples. YWHAQ was detected in only a small subset of samples and failed quality-control filtering, whereas YWHAH, YWHAB, YWHAG, and SFN were not detected. Thus, the serum proteomic data do not directly assess YWHAH, the paralogue most specifically implicated in reported Na_v_1.5 interactions. Four peptides mapping to YWHAE were identified; however, none was specific to a single YWHAE isoform. The data therefore cannot distinguish canonical from alternative YWHAE isoform abundance or establish an isoform switch (Supplementary table S1).

Paired-change analyses did not reveal a uniform co-change pattern among YWHAE, YWHAZ, MMP9, and CTSG; none of the six candidate-candidate correlations reached nominal P<0.05 in either sampling compartment. Platelet-associated markers PF4V1 and PPBP were evaluated. Direct platelet-marker changes did not consistently correlate with YWHAE, YWHAZ, or CTSG. The strongest platelet-related associations involved MMP9: arterial ΔMMP9 correlated with ΔPF4V1 (Spearman rho=0.69, P=0.012); the coronary-sinus PPBP-MMP9 association was weaker (rho=0.53, P=0.057) (Figure S4). Neutrophil-marker associations were likewise compartment dependent: arterial MPO correlated with YWHAZ (rho=0.64, P=0.021), arterial LTF with YWHAE (rho=0.59, P=0.023), coronary-sinus MPO inversely with CTSG (rho=-0.62, P=0.037), and coronary-sinus LTF with MMP9 (rho=0.70, P=0.0069) (Figure S5).

The paired-sample heatmap demonstrated heterogeneous abundance patterns across patients and between arterial and coronary-sinus compartments. AF-SR differences were not visually uniform across YWHAE, YWHAZ, or the broader platelet- and neutrophil-associated marker panels (Figure S6).

### AF Serum Modestly Reduces Ca²⁺-Transient Amplitude Without Increasing Diastolic Ca²⁺ Release

Ca²⁺ handling was assessed in iPSC-aCMs exposed to paired SR and AF serum from seven patients. Representative line-scan recordings demonstrated paced Ca²⁺ transients and spontaneous diastolic Ca²⁺-release events under both serum conditions (Figure 6A and 6B). AF-serum exposure was associated with a modest but statistically significant reduction in Ca²⁺-transient amplitude compared with paired SR-serum exposure (P<0.05; Figure 6C). In contrast, time to 50% release, time to peak, and the monoexponential decay constant (Tau) did not differ significantly between the two serum conditions (Figure 6D through 6F). Spontaneous Ca²⁺-spark frequency was also comparable after SR- and AF-serum exposure (Figure 6G). Thus, under the experimental conditions used, AF serum attenuated the amplitude of paced Ca²⁺ release but did not accelerate Ca²⁺-transient kinetics or increase spontaneous diastolic sarcoplasmic-reticulum Ca²⁺ release.

**Figure 6.**
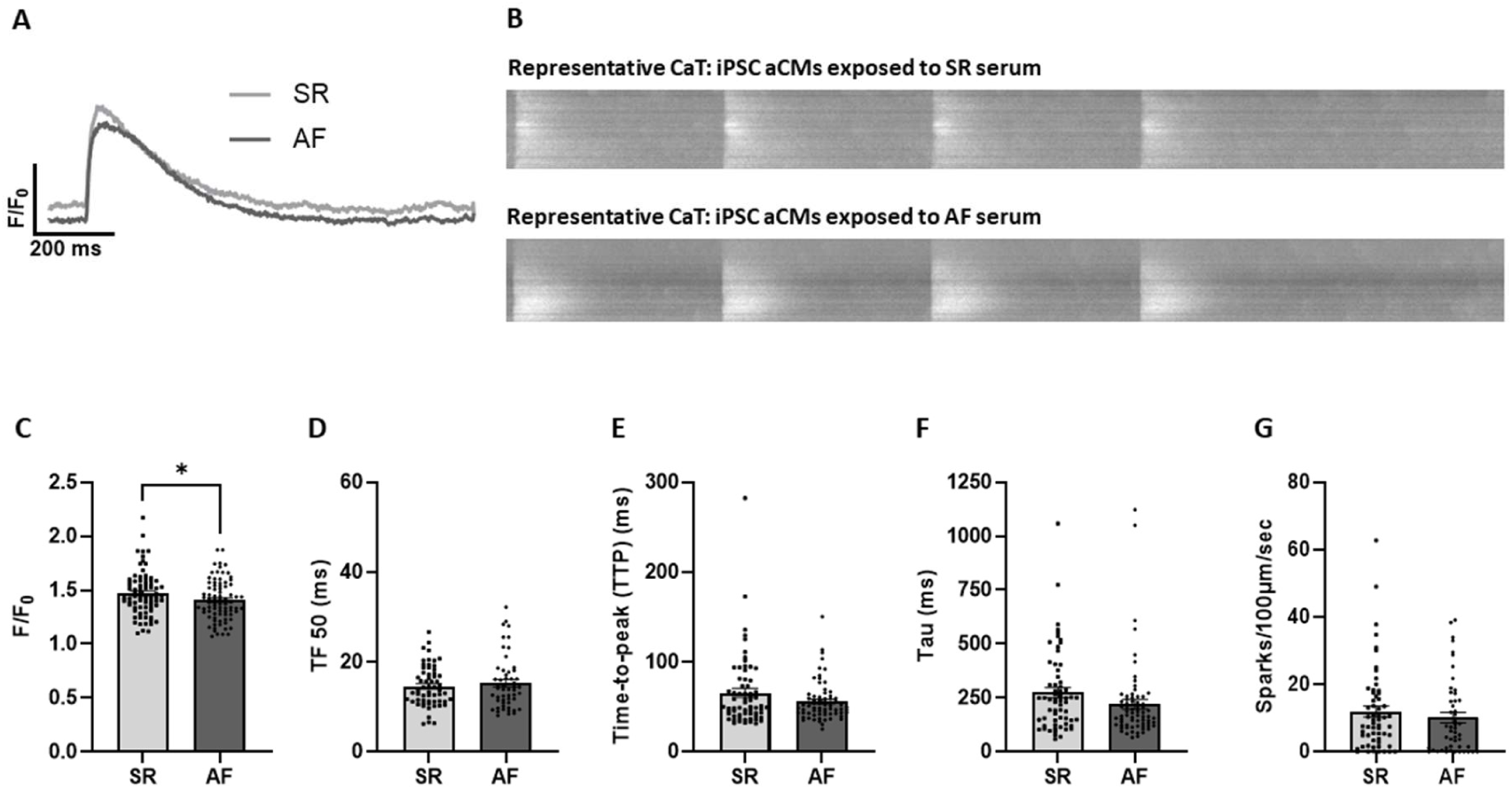
Altered calcium handling in iPSC derived atrial cardiomyocytes exposed to patient serum. **(A)** Representative Ca²⁺ transient traces (F/F₀) recorded during 1 Hz pacing in cells treated with serum from patients in sinus rhythm (SR, light grey) or atrial fibrillation (AF, dark grey). **(B)** Representative line-scan images of Ca²⁺ transients acquired during 1 Hz stimulation under SR and AF conditions. **(C–G)** Quantitative analysis of Ca²⁺ transient characteristics derived from line-scan recordings (*n* = 7 patient samples). **(C)** Average Ca²⁺ transient amplitude (F/F₀). **(D)** Average Time to 50% decay (TF50). **(E)** Average Time-to-peak (TTP). **(F)** Average Decay time constant (Tau). **(G)** Average Ca²⁺ spark frequency (sparks·100 µm⁻¹·s⁻¹). Individual data points represent single cells; bars indicate mean ± SD. Statistical comparisons were performed using paired Student’s *t*-test; *p*-values are indicated where significant.

### Differential modulation of SR serum–induced Ca²⁺ dysregulation by targeted inhibitors

To test whether candidate proteins identified by proteomics contribute to serum-induced Ca²⁺ dysregulation in iPSC-aCMs, pharmacological inhibition of MMP9, 14-3-3 signaling (Difopein), or Cathepsin G was performed during SR serum exposure. While Ca²⁺ transient amplitude remained preserved across all conditions, inhibition of MMP9 modestly impaired Ca²⁺ relaxation, prolonging TF₅₀ and τ, and markedly increased spontaneous Ca²⁺ spark frequency, consistent with exacerbated diastolic SR Ca²⁺ leak. In contrast, inhibition of 14-3-3 signaling exerted only modest effects on Ca²⁺ kinetics and did not further increase spontaneous Ca²⁺ release. Cathepsin G inhibition tend to accelerate the onset of Ca^2+^ release, and shortened Ca²⁺ transient duration without increasing spark frequency, suggesting a stabilizing effect on diastolic SR Ca²⁺ handling (Figure 7).

**Figure 7.**
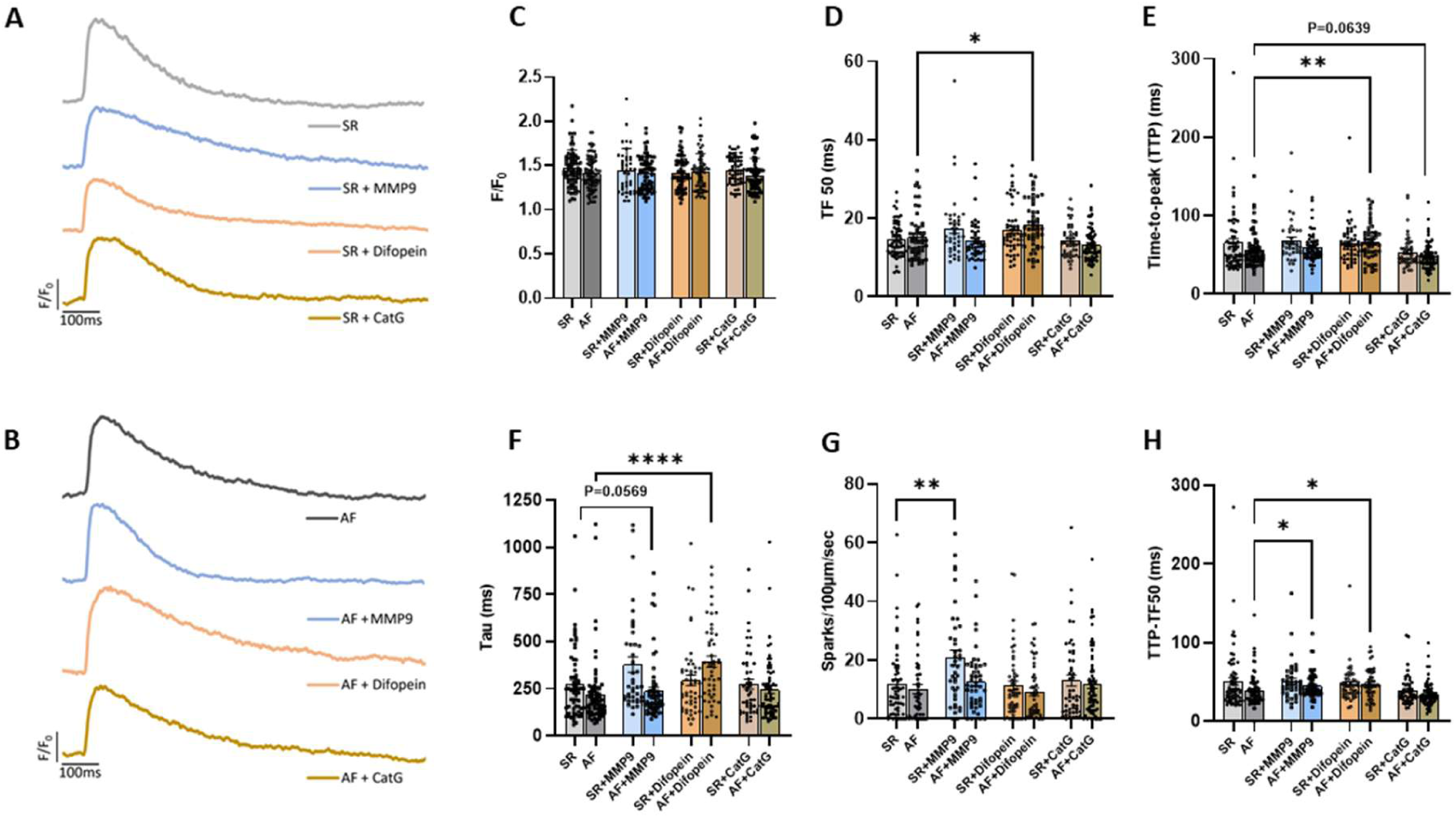
Distinct modulation of patient ‘s serum by targeted inhibitors on Ca²⁺ handling in human iPSC-derived atrial cardiomyocytes (iPSC-aCMs). **(A-B)** Representative normalized Ca²⁺ transient traces (F/F₀) recorded from iPSC-aCMs exposed to serum from N=7 patients alone or in the presence of pathway inhibitors: matrix metalloproteinase-9 (MMP9), 14-3-3 (Difopein), and cathepsin G (CatG) (A-SR serum, B-AF Serum). Traces are time-aligned; scale bar, 100 ms. **(C–H)** Quantification of Ca²⁺ transient properties: (C) Average peak amplitude (F/F₀), (D) Average time to 50% release (TF50), and (E) Average time-to-peak (TTP), (F) Average decay time constant (Tau), (G) Ca²⁺ spark frequency (sparks per 100 μm per s), and (H) Diastolic Ca2+ kinetics (TTP–TF50) cross all panels, iPSC-aCMs were incubated with patient serum (N=7 independent samples), and measurements represent pooled single-cell analyses from 7 independent AF serum samples tested. Statistical comparisons were performed using one-way ANOVA with post hoc testing as appropriate; exact P values are indicated where significant.

### Distinct effects of inhibitors on AF serum–induced Ca²⁺ abnormalities

In AF serum–treated cardiomyocytes, Ca²⁺ transient amplitude was preserved under all inhibitor conditions. MMP9 inhibition did not significantly alter Ca²⁺ kinetics or spontaneous Ca²⁺ spark frequency compared with AF serum alone. In contrast, inhibition of 14-3-3 signaling markedly slowed Ca²⁺ removal, with prolonged TF₅₀ and τ, accompanied by a modest reduction in spontaneous Ca²⁺ spark activity. Cathepsin G inhibition further accelerated Ca²⁺ release kinetics while maintaining stable diastolic Ca²⁺ release (Figure 7). These data indicate that distinct circulating factors contribute to Ca²⁺ dysregulation in SR and AF serum, with context-dependent effects on intracellular Ca²⁺ homeostasis.

### Exploratory Associations with Atrial Disease Severity

Patient-specific Ca²⁺-transient amplitudes were related to clinical and electroanatomic measures of atrial remodeling. No statistically significant association was identified with left atrial volume index (A, rₛ=-0.085, P=0.807), low-voltage area burden (B, rₛ=-0.717, P=0.178), left atrial pressure (C, rₛ=-0.443, P=0.211), or log-transformed NT-proBNP (D, rₛ=0.537, P=0.078; Figure 8). The inverse effect estimates for low-voltage area and left atrial pressure and the positive effect estimate for NT-proBNP were hypothesis-generating but imprecise because of the limited number of patients with complete paired experimental and clinical data. Supplemental analyses relating integrated sodium current to low-voltage burden similarly did not identify statistically significant associations (Figures S1 through S3).

**Figure 8.**
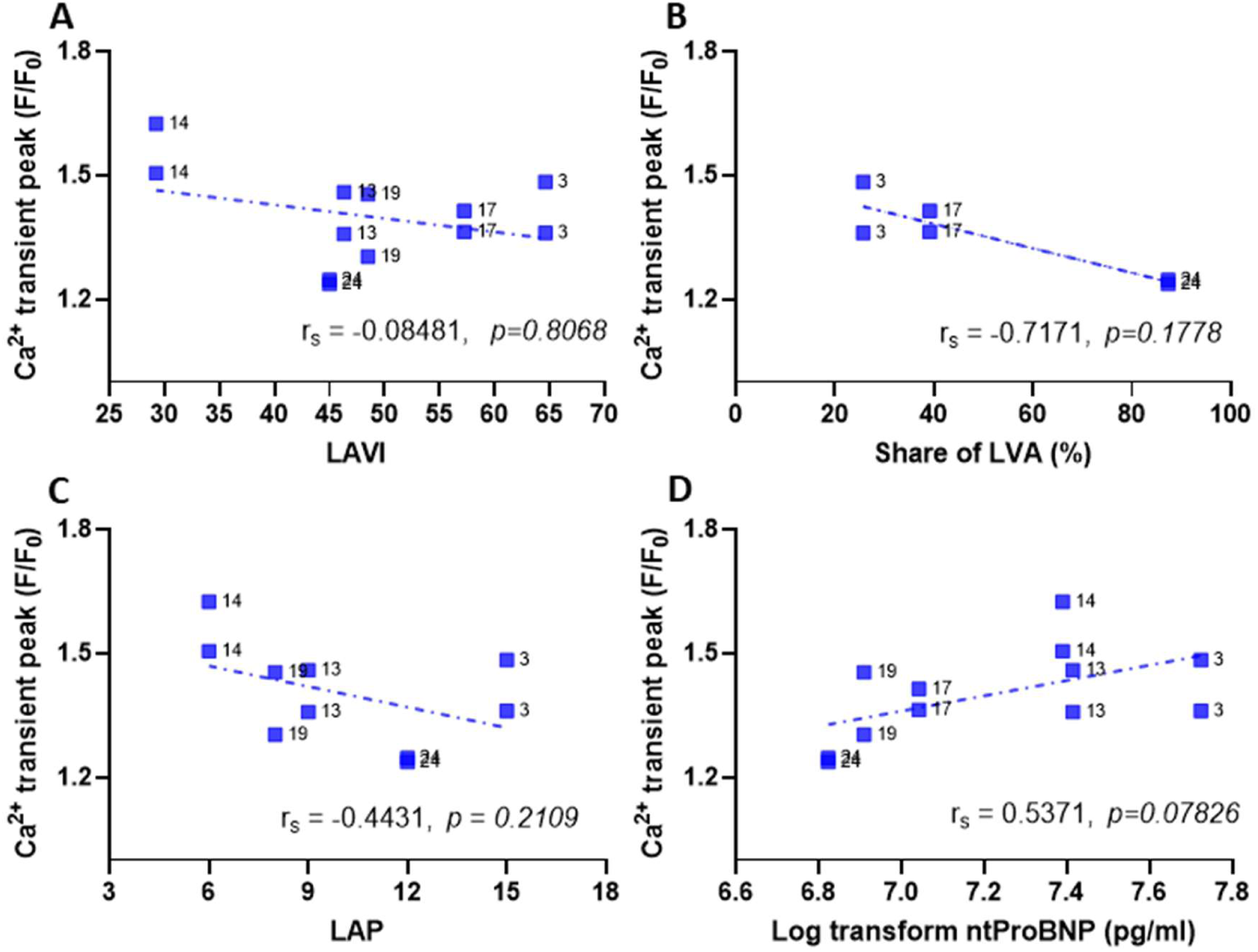
In vitro – In vivo Correlation between patient-specific mean Ca²⁺ transient amplitude (F/F0) and individual in-vivo measures. Correlations between Ca^2+^ transient peak (F/F₀) and (**A**) left atrial volume index (LAVI), (**B**) percentage of low-voltage area (Share of LVA), (**C**) left atrial pressure (LAP), and (**D**) log-transformed N-terminal pro–B-type natriuretic peptide (nt-proBNP). Statistical relationships were assessed using Spearman rank correlation and simple linear regression model.

## Discussion

This study provides evidence that serum from patients with persistent AF has transferable electrophysiological bioactivity. In an isolated-heart preparation, AF serum produced the greatest duration and cumulative burden of spontaneous atrial depolarizations, whereas serum obtained after restoration of SR showed an intermediate phenotype. At the cellular level, AF serum caused more selective effects, including a reduction in Ca^2+^ transient amplitude and limited modulation of sodium-current properties. Proteomic profiling further suggested that the circulating phenotype is multifactorial and rhythm-context dependent rather than attributable to a single dominant mediator.

The principal conceptual advance is the functional transfer of a patient-derived circulating phenotype into an intact cardiac preparation. The paradigm that “AF begets AF” was established by the observation that sustained AF promotes electrical remodeling and increasing arrhythmia stability. (1) Contemporary concepts extend this framework to atrial cardiomyopathy, in which fibrosis, inflammation, oxidative stress, and electrical remodeling interact to sustain AF (11, 12) which aligns with guideline-level views of AF as a chronic, multifactorial disease. (13)

Most circulating biomarker and proteomic studies have treated blood proteins as correlates of AF presence, recurrence, or substrate severity.(6–8) The present results suggest that the circulation may also participate directly in arrhythmogenic signaling.

The disparity between the prominent intact-heart phenotype and the comparatively modest effects in isolated cardiomyocytes is mechanistically informative: Atrial electrical instability emerges from the interaction of cellular electrophysiology with tissue architecture, conduction reserve, cell-cell coupling, fibroblast and endothelial signaling, and extracellular matrix. Small changes across several pathways may therefore be amplified in intact tissue even when no single cellular endpoint is markedly altered. Conversely, human induced pluripotent stem cell-derived atrial cardiomyocytes remain developmentally immature and cannot reproduce the multicellular substrate of advanced atrial cardiomyopathy. The present data should therefore not be interpreted as evidence that AF serum reproduces the canonical Ca^2+^-leak phenotype of chronically remodeled human atria. Enhanced diastolic sarcoplasmic-reticulum Ca^2+^ release is a recognized driver of delayed afterdepolarizations in AF (2, 3) and broader mechanistic models link Ca²⁺ dysregulation to both triggers and remodeling in AF. (14–17) Even though AF serum did not consistently increase Ca^2+^ spark frequency in this study, it decreased Ca^2+^ transient amplitudes potentially reflecting altered Ca^2+^ entry, sarcoplasmic-reticulum loading, release gain, or adaptive Ca^2+^-signaling silencing, a process described in sustained AF. (18, 19)

The proteomic and inhibitor experiments support a network-based interpretation. MMP9 is linked to extracellular-matrix turnover and AF recurrence (20, 21), whereas 14-3-3 proteins regulate phosphorylation-dependent trafficking and signaling of cardiac ion-channel complexes. However, the biological interpretation of these circulating candidates requires caution because part of their variation may reflect blood-cell– or sample-processing–related effects rather than rhythm-specific cardiac release. In exploratory paired analyses, changes in MMP9 correlated with platelet-associated PF4V1 in arterial samples and with the neutrophil-associated marker LTF in coronary-sinus samples, while the association with platelet-derived PPBP in coronary-sinus serum was weaker. These relationships indicate that platelet activation, coagulation-related processes, and neutrophil activation may contribute to the observed MMP9 signal and therefore limit its interpretation as a specific AF-derived mediator. Similarly, associations between YWHAE and markers potentially reflecting blood-cell contamination or hemolysis raise the possibility that at least part of the circulating YWHAE signal may be influenced by preanalytical variation rather than AF biology alone. These findings do not exclude a biological role for either MMP9 or YWHAE, but they emphasize that their cellular origin cannot be inferred from serum proteomics and that targeted validation in carefully controlled samples will be required.

The 14-3-3 findings also require consideration of paralogue and isoform specificity. The rationale for examining YWHAE peptides arose from the potential relationship between 14-3-3 proteins and cardiac sodium-channel regulation, particularly Na_v_1.5. However, among the seven human 14-3-3 paralogues, only YWHAE and YWHAZ were robustly quantified, whereas YWHAH—the paralogue most specifically implicated in reported Na_v_1.5 interactions—was not detected in the present dataset. Furthermore, although four YWHAE-derived peptides were identified, none uniquely distinguished the YWHAE isoform. The proteomic data therefore cannot establish whether the rhythm-associated YWHAE signal reflects a particular isoform or an isoform switch. Accordingly, the relationship between the serum 14-3-3 signal and the observed sodium-current phenotype remains mechanistically suggestive rather than directly demonstrated. Moreover, disruption of 14-3-3 interactions mainly altered Ca^2+^ transient kinetics under AF-serum conditions, while e.g. Cathepsin G inhibition produced no consistent rescue. These findings argue against a simple one-protein/one-phenotype mechanism. Instead, rhythm-dependent serum bioactivity is likely generated by interacting proteolytic, inflammatory, and signaling pathways whose effects depend on the prevailing extracellular and cellular context.

The exploratory relationships between serum-induced cellular responses and low-voltage burden, left atrial pressure, and NT-proBNP are potentially relevant to atrial cardiomyopathy but remain inconclusive. Although low-voltage areas identify diseased atrial substrate and are associated with AF recurrence after ablation (5, 22, 23), serum-induced cellular responses were not significantly associated with low-voltage burden, left atrial pressure, or NT-proBNP in the present cohort. This suggests that the measured circulating bioactivity does not simply mirror the severity of underlying atrial cardiomyopathy.

Several limitations warrant emphasis. The cohort and functional subsets were small, and SR serum was collected 15 minutes after cardioversion; thus, these samples represent an early post-cardioversion state rather than stable SR. Acute effects of cardioversion, anesthesia, anticoagulation, and procedural hemodynamics cannot be excluded. The Langendorff experiments used a fixed control-SR-AF sequence, leaving potential time and carryover effects despite washout. The isolated-heart model was cross-species, and the spontaneous-depolarization metric is an experimental index of atrial instability rather than direct documentation of clinical AF. Cellular experiments used a single iPSC line and separate cells for the serum conditions, requiring patient-serum-level rather than cell-level inference. Finally, proteomic candidates were nominated by association and pharmacological perturbation but were not confirmed by depletion/add-back experiments.

In conclusion, patient serum exerts transferable electrophysiological effects, with the strongest intact-heart bioactivity observed during AF. The accompanying cellular phenotype is selective and does not simply reproduce chronic tissue-intrinsic AF remodeling. These findings extend the “AF begets AF” concept by identifying the circulation as a potential, and potentially reversible, contributor to atrial electrical instability.

This project was funded by the following grants: Deutsche Forschungsgemeinschaft (German Research Foundation; SFB-1470, B01) and the Investitionsbank Berlin (co-funded by the EU; ENABLED-Heart).

## Data Availability

All the data referred in this manuscript is available

---

**Supplementary Table S1.**
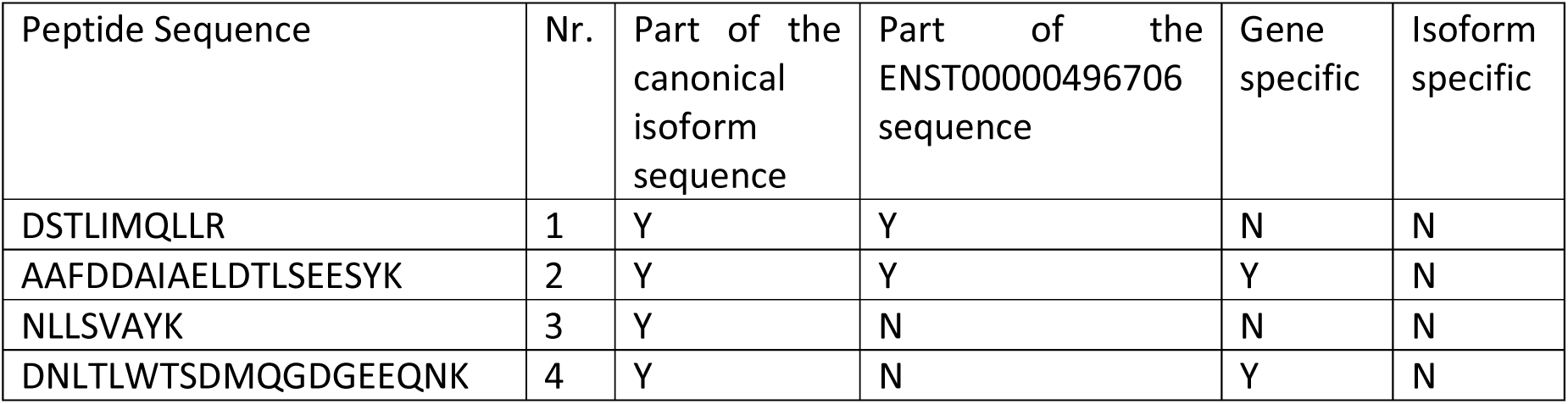
YWHAE peptide sequence mapping and assessment of isoform specificity.

